# Enormity of anaemia and its determinant factors among lactating mothers in Northern Ghana: A case of nanton district

**DOI:** 10.1101/2022.06.18.22276586

**Authors:** Adadow Yidaana, Enoch Weikem Weyori, Khadijah Wemah, Nkrumah Bernard, Braimah Baba Abubakari, Kuugbee Dogkotenge Eugene, Benjamin Nuertey, Etowi Boye Yakubu, Valentine Koyiri Cheba, Adatsi Richard Kujo

## Abstract

**Background:** Anemia remains one of the most severe and common public health conditions that predominantly affects children and women across the globe. It is defined as a condition in which the hemoglobin (Hb) concentration is less than 11.0 g/dl particularly in women. The World health organization report indicated that 20–50% of the world population was affected by iron deficiency anemia.

**Method:** An institutional cross-sectional study design was the method used through the data collection and management. Information was sought from four selected health centres across the nanton district with systematic sampling deployed to select respondents of interest. A sample of 420 respondents were obtained and processed for analysis. A bi-variate and multivariate analysis uncovered the associated factors and its predictiveness.

**Results:** The prevalence of anaemia in totality was 56.0% (95% CI 51.3%, 60.8%). The divergence of the blood concentration levels between the means of the anaemic and non anaemic group was statistically significance (**F-stat**=68.233, **t-stat**=-35.697, **p**<0.01). The multivariate statistical model showed that, lactating mothers who have suffered malaria episode(s) after delivery had a 94% chance of being anaemic [**AOR** = 0.054; (95% **CI**: 0.025, 0.119)]. Lack of post-partum iron supplementation increased the risk of having anaemia [**AOR** = 15.336; (95% **CI**: 6.112, 38.483)], and lactating mothers had higher risk [**AOR** = 1.927, (95% **CI**: 1.031, 3.602)] of developing anaemia with increasing ‘child’s age beyond three (3) months.

**Conclusion:** Anaemia remains very high in lactating mothers attributable to episodes of postpartum malaria, iron supplementation, and increasing ‘child’s age beyond 3 months. There is the need for public health interventions and measures such as extension of folic acid distribution and Intermittent Preventive Therapy (IPT) for malaria program to mothers at postnatal care and child welfare clinics across the region.

## Introduction

Anaemia is a deficiency of red cells or haemoglobin in the blood, resulting in pallor and weariness. In developing countries, anaemia remains a pertinent dietary issue and a crucial challenge for human’ ‘ health, as well as social and economic progress [3]. Anemia is still more common and widespread in developing countries than in industrialized countries, with Southeast Asia and Sub-Saharan Africa bearing the brunt of the disease’s burden [4-6]. Worldwide, an estimated 54.1% of anemia cases contribute to mild, 42.5% moderate, and 3.4% severe problems in all age groups of the population [7, 8]. The estimated burden of anaemia and its magnitude amongst lactating mothers is 52.5% in South Asia, 60.3% in Myanmar, 20% in Nepal, 63% in India, and 28.3% in Ethiopia [6, 9-11].

In 2015 and 2016, the Indian National Family Health Survey (NHFS-4) revealed that 53% of women in India are anemic. Mild anemia affects 40% of women, moderate anemia affects 12% of women, and severe anemia affects 1% of women. Anemia affected 58% of breastfeeding women, with 44.5% having a mild case, 12.6% having a moderate case, and 0.9% having a severe case [12].

According to the Ghana Statistical Service (2014), 42% of women in Ghana are anaemic, 32% were mildly anaemic, 10.0% were moderately anaemic, and less than 1% were severely anaemic. The highest prevalence of anaemia is among the youngest women aged 15-19 (48%), and the least prevalence was recorded among women age 30-39 (39%) [13]. Out of 1,041 lactating mothers 45% were anaemic; 39% mild, 8.1% moderate and 0.1% severe. Anaemia prevalence among pregnant women was closer to that among lactating women (45%) but slightly higher than the prevalence among women who are neither pregnant nor breastfeeding (41%). Residency did not make a crucial distinction on the prevalence of anaemia among women [13].

Anemia is more common among lactating mothers in the poor wealth index group, the unemployed, mothers breastfeeding for the last year, high illiteracy rate, BMI ≥ 25 kg/m^2^, high parity, and no iron pill supplementation during pregnancy [14 – 18].

Anemia is a multifaceted disorder with numerous causes. Low intake of absorbable dietary iron to fulfill the body’s demands, especially during lactation and pregnancy, as well as during periods of fast growth in children, causes anaemia. Studies conducted in Ghana have revealed that malaria and helminthic infections, iron deficiency, vitamin A deficiency, ‘mother’s educational status, and wealth quintiles are major contributors of anaemia in lactating mothers [19 – 21]. The unchanging situation with respect to anaemia in pregnancy and limited findings of anaemia in lactating mothers has driven the need to investigate the current situation of anaemia prevalence and its contributing factors in lactating mothers in the northern region of Ghana.

## Methodology

### Study Setting

The study area included communities in the four sub-districts of the Nanton district. The sub-districts are Nanton, Zoggu, Tampion, and Janjori-Kukuo.

### Research Design

A cross-sectional survey was the design for this study. The cross-sectional study design helped in setting out the most appropriate framework to capture relevant information needed to accomplish this study.

### Study Population

The targeted population for this study was comprised of all postpartum mothers who delivered within the last 6 months and in attendance to Child Welfare Clinics within the study period.

### Sample Frame

The child welfare clinic register was introduced as the sampling frame for selecting the respondents who participated in the study across all selected and captured health facilities within the district.

### Study subjects

The study subjects included all postpartum mothers who delivered within the last six months and attended CWC in the study health facilities within the study period and who consented to participate in the study.

### Sample Size Calculation

The sample size was calculated based on the population of Child Welfare Clinic (CWC) of the Nanton district over the 2020 period. The sample calculator that best suits the population was the single population proportion (SPP) formula where;

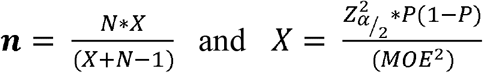

**n** - Represents the required sample

**P** – Probability of a postpartum mothers to attend PNC services in a health facility

**(1-P)** – Probability of a postpartum mothers not attending PNC services in a health facility

**N** - Represents the population estimated postpartum mothers in the Nanton district for 2020

**MOE** – is the allowable margin of error set for the study by the researcher

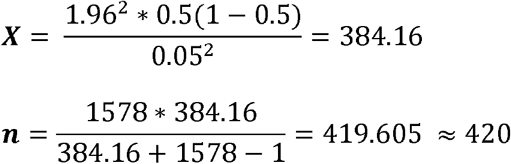

### Sampling Technique

The systematic sampling procedure was adopted and deployed for the selection of respondents to participate in the study. Lactating mothers were selected using the systematic approach to sample from the sample frame (CWC register) of the respective health centres of the Nanton district. This process of selection was done for twenty days using trained field personnel. The sampling frame was used as a proxy for the selection of respondents who participated in the study.

### Measurement of haemoglobin

To measure the dependent variable of interest, the haemoglobin of all respondents was measured using a haemoglobinometer (*brand of haemoglobinometer and manufacturer and any other describable features of the device is needed*) during the study. The haemoglobinometer uses spectrophotometric measurement to determine the value of the hemoglobin levels. Capillary skin puncture was performed to obtain blood sample from the three middle finders of the lactating mothers using the prick and the micropipette and transferred into the microcuvette for measurement. The microcuvette with the sample is placed in the Haemocue HB 201 reader. Results were obtained by a trained laboratory professional who assisted in the process.

### Data analysis and Presentation

Data was collected through an institutional cross-sectional design which included selected variables in determining the related associations and significance levels. Minitab statistical software was key in analysis and outcomes of the findings. Descriptive statistical outcomes were presented in the forms of frequency tables, graphs, and charts for easy understanding and interpretation. The prevalence was calculated and subjected to significant testing for statistical significance.

T-test statistic for small samples was applied to check the significant difference between the average HB of anaemic mothers and that of non-anaemic mothers. Also, Bi-variable and multi-variable effect analysis was conducted on anaemia status among lactating mothers to determine the predictive factors associated with anaemia. A p-value cut-off point for significance was set at p<0.05, representing the significance level of each factor.

## Findings and Results

### Socio-demographic characteristics of the study participants

The study presented a total weighted sample of 420 participants from the Nanton district of the Northern region, with a larger proportion not having any form of formal education. All participants of the study were employed in one way or the other, with the majority being married. The majority of lactating mothers (199; 47.4%) included in the study were in the age range of 30–39 years. Almost all lactating mothers (96.4%) were Muslims, with a greater proportion (94.8%) being dagomba’s. Age category (p<0.001), ethnicity (p=0.043), and educational status (p<0.001) of lactating mothers had a statistically significant association with the anaemia status (**Table 1**).

**Table 1:**
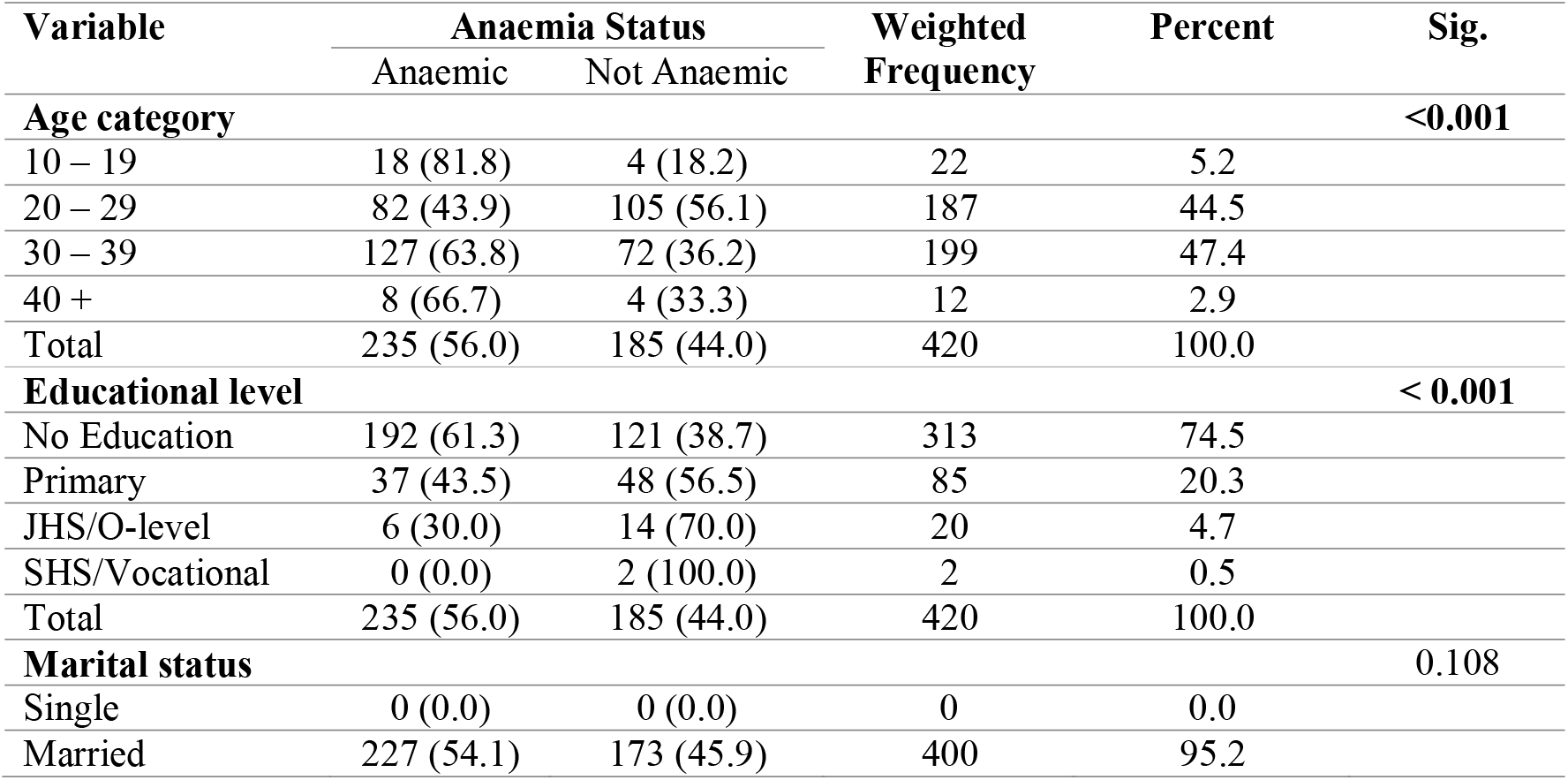
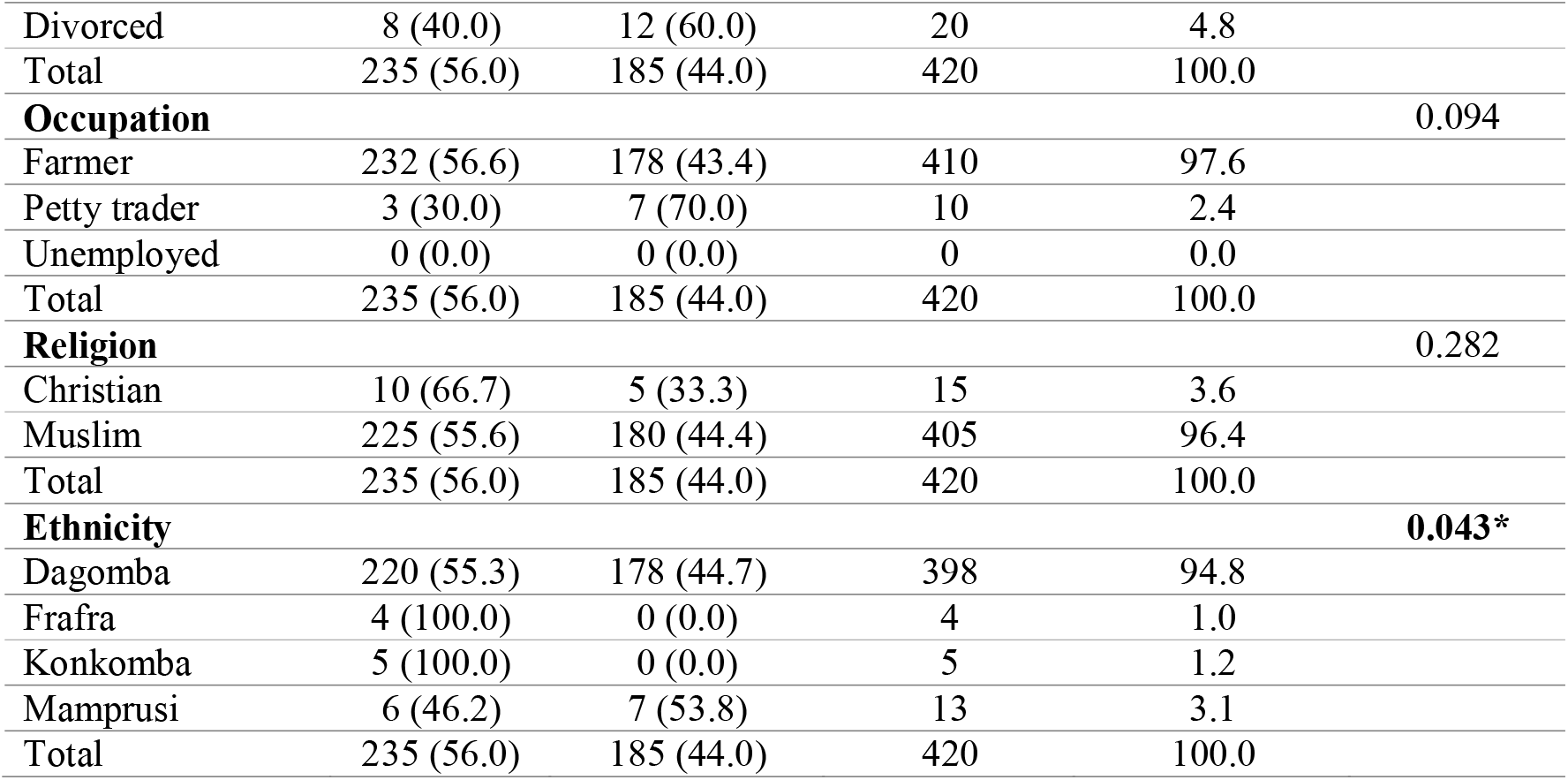
Socio-demographic characteristics

### Behavioral and Obstetric Factors

Outcomes depict that 56.7% of lactating mothers have breastfed their children exclusively within one to three months, most lactating mothers (84.3%) were multi-parous, and a greater proportion (96.4) had given birth by spontaneous vaginal delivery (SVD). Contraceptive usage (12.6%) was generally low amongst lactating mothers but more predominant amongst mothers who were anaemic (41; 88.1%). Around 43.3% of mothers include complementary feeding, only 6.9% of the lactating mothers use long-lasting insecticide-treated nets, and 32.4% had treated one or more episodes of malaria within the lactating period. Iron supplement usage (16.0%) was very low, the majority of the lactating mothers (98.1%) did not deworm after birth, and a greater proportion (54.8%) of the children were four and above. Again, parity, malaria, and iron supplementation were significant predictors of anaemia amongst lactating mothers (Table 2)

**Table 2:**
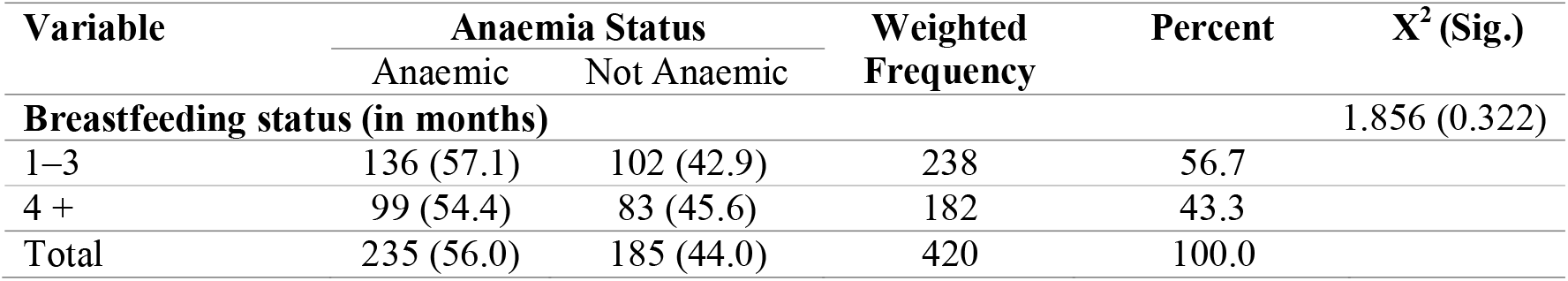
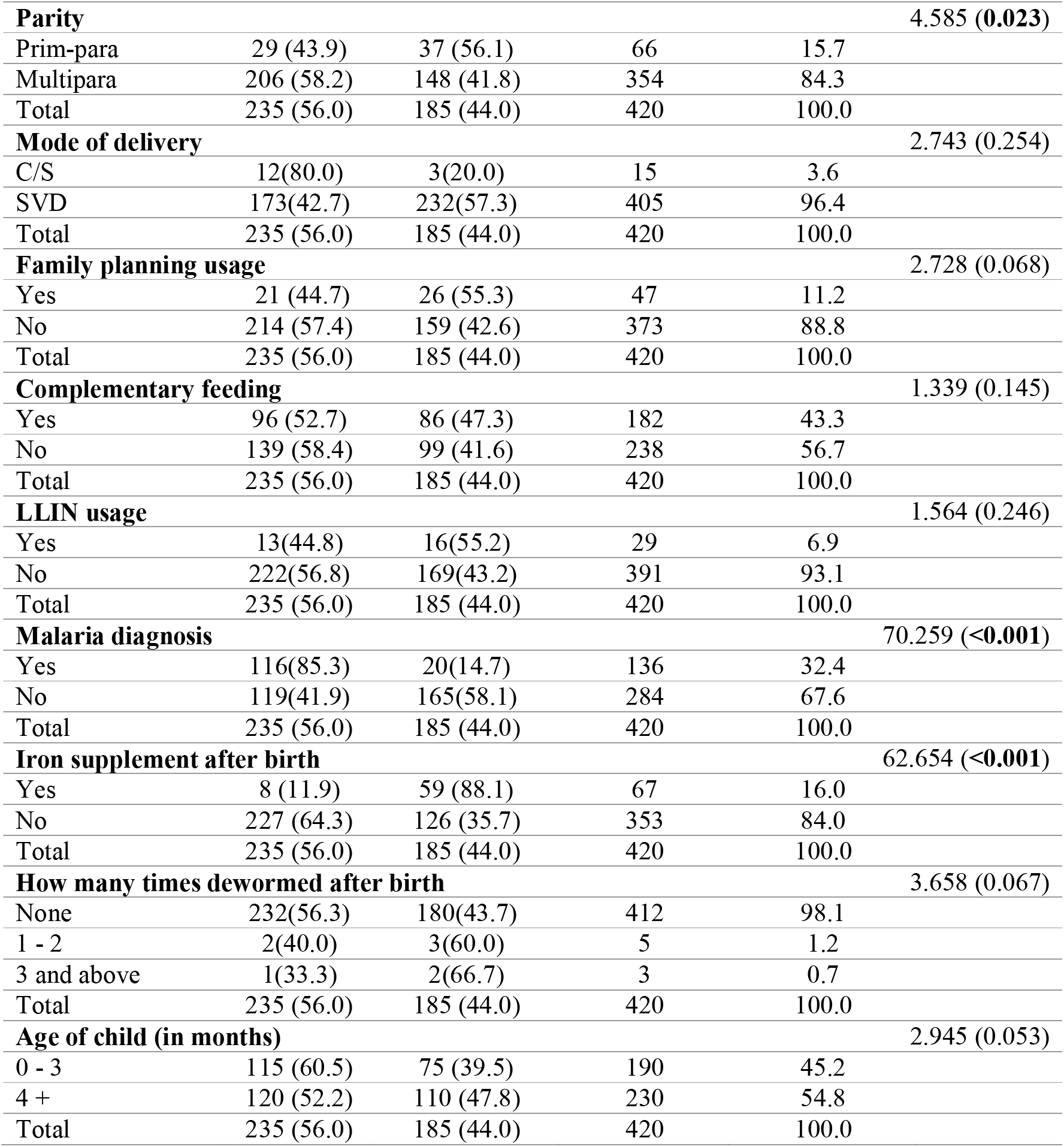
Behavioral and obstetric characteristics of study participants

### Magnitude of anaemia amongst lactating mothers in Nanton

The overall prevalence of anemia estimated among lactating mothers in Nanton was 56.0% [95% confidence interval (CI): 51.25, 60.75%]. With respect to the anaemia severity of the lactating mothers, 18.7% reported had mild anaemia; 67.20% had moderate anaemia, and 14.0% of the mothers had severe anaemia, respectively (Figure 1). Proportionately anaemia prevalence was highest among lactating mothers (81.8%) in the age group of less than 20 years and lowest (56.10%) in the age group of 20–29 years (Figure 1).

**Figure 1:**
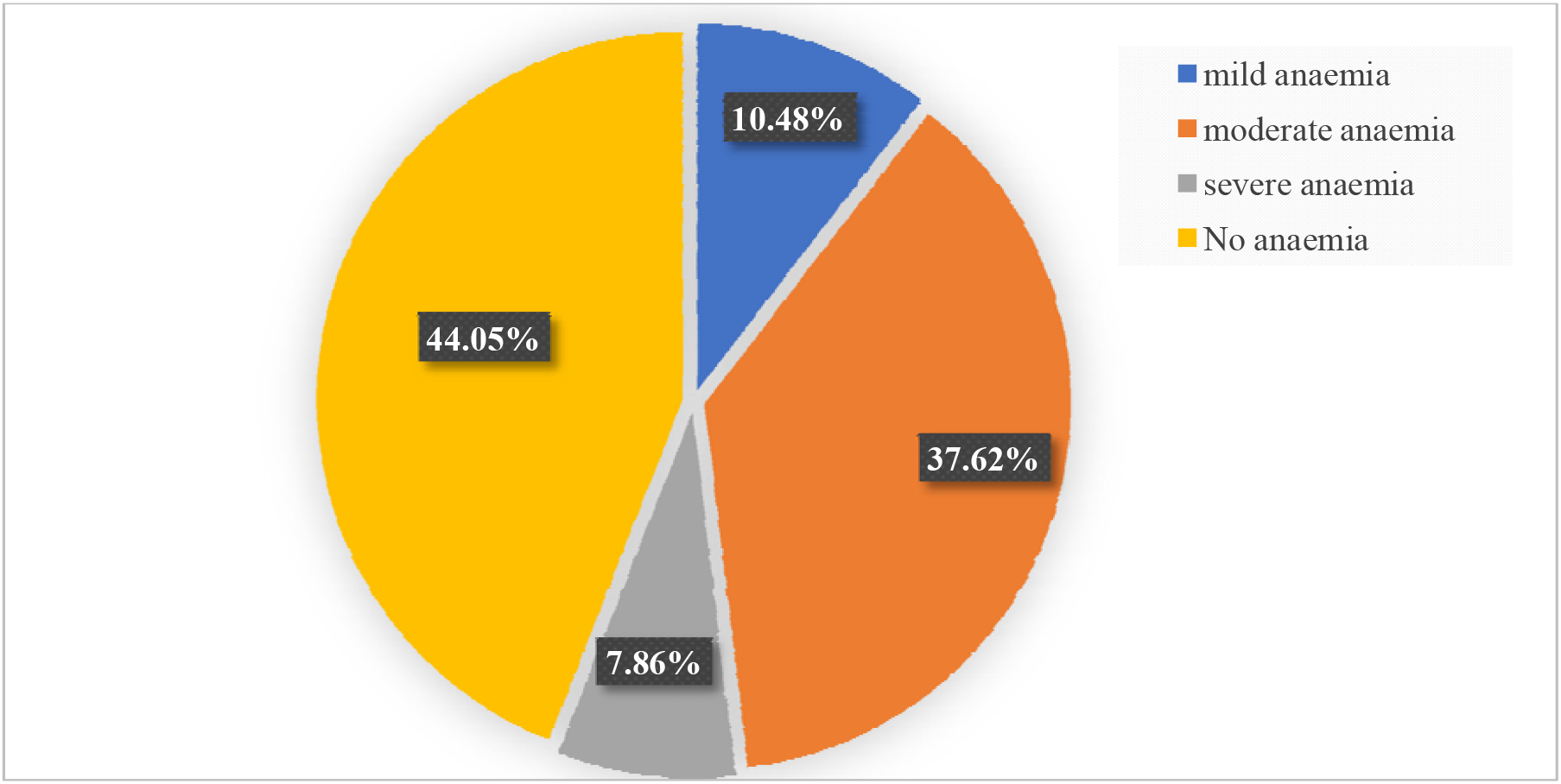
Magnitude of anaemia amongst lactating mothers in Nanton District

### Divergence in the mean of anaemic and non-anaemic mothers

Table 3, the average mean of anaemic mothers was 8.7081(SD=1.35157, SE=0.08817) compared to mothers who are not anaemic with a mean of 12.4530 (SD=0.77327, SE=0.05685). There was a statistically significant difference between haemoglobin levels of anaemic mothers and that of non-anaemic mothers (F=68.233, MD=3.74489, p<0.001).

**Table 3:**
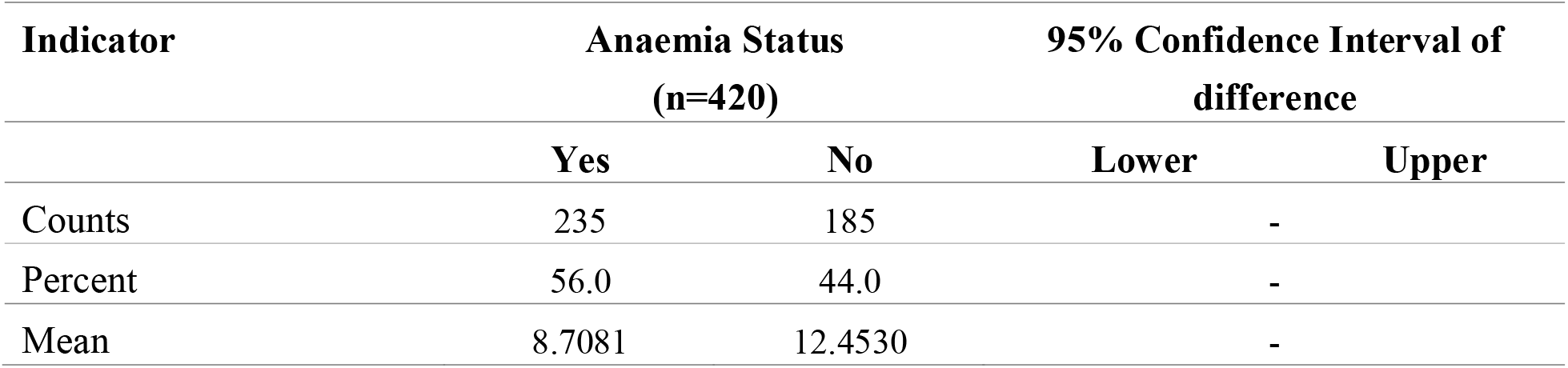
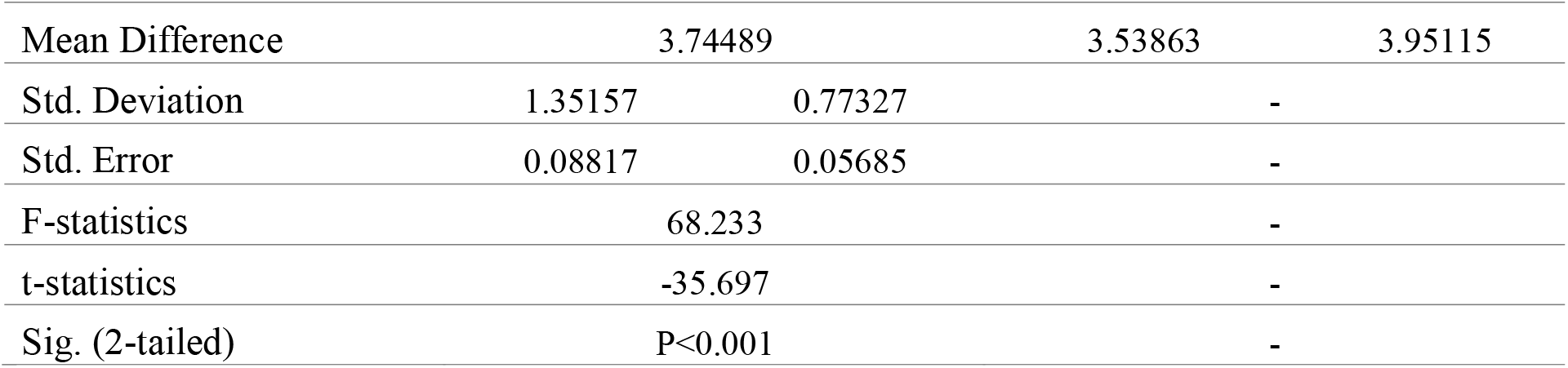
Difference in anaemia status

### Predictive factors associated with the anaemia status of lactating mothers

The outcomes revealed malaria diagnosed after birth, iron supplementation, and age group are predictive factors of anaemia among lactating mothers.

The odds of having anemia increased by 94.6% amongst lactating mothers who have had episode(s) of malaria after birth (AOR = 0.054, 95% CI: 0.025, 0.119) compared to lactating mothers who did not have any episode(s) of malaria. Similarly, the odds of having anaemia in lactating mothers who do not take iron supplements is 15 times (AOR = 15.336, 95% CI: 6.112, 38.483) more than a lactating mother who takes iron supplements. Lactating mothers had a higher risk (AOR = 1.927, 95% CI: 1.031, 3.602) of developing anaemia with an increasing ‘child’s age greater than three (3) months compared to lactating mothers with children below four (4) months (shown in Table 4).

**Table 4:** Bi-variable and multi-variable effect analysis of anemia among lactating mothers

| Variable | Anaemia Status |  | Odds Ratio [95% CI] |  | P-Value |
| --- | --- | --- | --- | --- | --- |
|  | Yes | No | COR | AOR |  |
| Breastfeeding status (in months) |  |  |  |  |  |
| 1–3 | 136 (57.1) | 102 (42.9) | 1 | 1 | - |
| 4 + | 99 (54.4) | 83 (45.6) | 1.118 [0.758-1.649] | 0.636 [0.103-3.933] | 0.626 |
| Parity |  |  |  |  |  |
| Prim-para | 29 (43.9) | 37 (56.1) | 1 | 1 | - |
| Multipara | 206 (58.2) | 148 (41.8) | 0.563 [0.331-0.957] | 1.289 [0.438-3.793] | 0.645 |
| Mode of delivery |  |  |  |  |  |
| C/S | 12 (80.0) | 3 (20.0) | 1 | 1 | - |
| SVD | 173 (42.7) | 232 (57.3) | 0.186 [0.052-0.671] | 5.897 [0.742-46.891] | 0.093 |
| Family planning usage |  |  |  |  |  |
| Yes | 21 (44.7) | 26 (55.3) | 1 | 1 | - |
| No | 214 (57.4) | 159 (42.6) | 1.666 [0.905-3.069] | 1.550 [0.651-3.691] | 0.322 |
| Complementary feeding |  |  |  |  |  |
| Yes | 96 (52.7) | 86 (47.3) | 1 | 1 | - |
| No | 139 (58.4) | 99 (41.6) | 1.258 [0.853-1.855] | 5.756 [0.868-38.189] | 0.070 |
| **LLIN usage** |  |  |  |  |  |
| Yes | 13(44.8) | 16(55.2) | 1 | 1 | - |
| No | 222(56.8) | 169(43.2) | 1.617 [0.757-3.453] | 2.963 [0.991-8.861] | 0.052 |
| **Malaria diagnosis** |  |  |  |  |  |
| Yes | 116(85.3) | 20(14.7) | 1 | 1 | - |
| No | 119(41.9) | 165(58.1) | 0.124 [0.073-0.211] | **0.054 [0.025-0.119]** | **<0.001** |
| **Iron supplement after birth** |  |  |  |  |  |
| Yes | 8 (11.9) | 59 (88.1) | 1 | 1 | - |
| No | 227 (64.3) | 126 (35.7) | 13.287 [6.153-28.692] | **15.336 [6.112-38.483]** | **<0.001** |
| **How many times dewormed after birth** |  |  |  |  |  |
| None | 232(56.3) | 180(43.7) | 1 | 1 | - |
| 1 - 2 | 2(40.0) | 3(60.0) | 1.933 [0.320-11.693] | 2.819 [0.423-18.804] | 0.285 |
| 3 and above | 1(33.3) | 2(66.7) | 2.578 [0.232-28.653] | 8.747 [0.455-168.199] | 0.151 |
| **Age of child (in months)** |  |  |  |  |  |
| 0 - 3 | 115 (60.5) | 75 (39.5) | 1 | 1 | - |
| 4 + | 120 (52.2) | 110 (47.8) | 1.406 [0.952-2.075] | **1.927 [1.031-3.602]** | **0.040** |
| **Age of mother** |  |  |  |  |  |
| 10 – 19 | 18 (81.8) | 4 (18.2) | 1 | 1 | - |
| 20 – 29 | 82 (43.9) | 105 (56.1) | 5.762 [1.878-17.682] | 4.725 [0.890-25.079] | 0.068 |
| 30 – 39 | 127 (63.8) | 72 (36.2) | 2.551 [0.831-7.829] | 2.385 [0.413-13.764] | 0.331 |
| 40 + | 8 (66.7) | 4 (33.3) | 2.250 [0.447-11.334] | 0.076 [0.002-3.545] | 0.189 |
| **Educational level** |  |  |  |  |  |
| No Education | 192 (61.3) | 121 (38.7) | 1 | 1 | - |
| Primary | 37 (43.5) | 48 (56.5) | 0.270 [0.101-0.722] | 0.198 [0.030-1.311] | 0.093 |
| JHS/O-level | 6 (30.0) | 14 (70.0) | 0.556 [0.195-1.586] | 0.333 [0.048-2.321] | 0.267 |
| SHS/Vocational | 0 (0.0) | 2 (100.0) | 6923 [6661-7230] | 4653 [3893-5734] | 0.899 |
| **Marital status** |  |  |  |  |  |
| Single | 0 (0.0) | 0 (0.0) | 1 | 1 | - |
| Married | 227 (54.1) | 173 (45.9) | 1.968 [0.787-4.920] | 0.964 [0.268-3.469] | 0.955 |
| Divorced | 8 (40.0) | 12 (60.0) | 0.508 [0.203-1.270] | 0.361 [0.221-1.109] | 0.967 |
| **Occupation** |  |  |  |  |  |
| Unemployed | 0 (0.0) | 0 (0.0) | 1 | 1 | - |
| Farmer | 232 (56.6) | 178 (43.4) | 3.041 [0.775-11.927] | 7.503 [0.391-143.991] | 0.181 |
| Petty trader | 3 (30.0) | 7 (70.0) | 0.329 [0.084-1.290] | 1.044 [0.091-3.098] | 0.121 |
| **Religion** |  |  |  |  |  |
| Muslim | 225 (55.6) | 180 (44.4) | 1 | 1 | - |
| Christian | 10 (66.7) | 5 (33.3) | 1.600 [0.537-4.765] | 2.019 [0.410-9.945] | 0.388 |
| **Ethnicity** |  |  |  |  |  |
| Dagomba | 220 (55.3) | 178 (44.7) | 1 | 1 | - |
| Frafra | 4 (100.0) | 0 (0.0) | 0.000 [0.00-0.00] | 0.000 [0.00-0.00] | 0.999 |
| Konkomba | 5 (100.0) | 0 (0.0) | 0.000 [0.00-0.00] | 0.000 [0.00-0.00] | 0.999 |
| Mamprusi | 6 (46.2) | 7 (53.8) | 1.442 [0.476-4.367] | 0.907 [0.210-3.928] | 0.517 |

*AOR, Adjusted Odd Ratio; COR, Crude Odd Ratio; CI, Confidence Interval. The bold values depict significant association with anaemia; 1, Reference category*

### Discussion of findings

Key public health concerns and interventions have overlooked the impact of anemia on lactating mothers and their children. Iron deficiency anaemia is the single most common micronutrient deficiency worldwide, affecting mothers and children [22]. Lactating mothers are more susceptible due to certain factors such as maternal iron depletion during lactation, blood loss during childbirth, and inadequate nutrient intake [1].

The study found the majority (74.5%) of mothers had no formal education, 95.2% were married, 96.4% were found to be Muslims, 15.7% of the children are first child of the mother, mothers recorded low malaria episodes (6.9%) after delivery and 384 (96.4%) delivered through spontaneous virginal delivery (SVD). Also uncovered was the mother’s age with a mean of 29.36 (SD=6.4) and the average number of children recorded to be 3.36 (SD=1.78). This finding is in divergence with the study conducted by Lakew et al. (2015) in Ethiopia found that most respondents (86%) were from rural settings, with nearly 60% of lactating mothers being Christians and 38% being Muslims, with a mean age 28.4 (SD=6.8) years [23]. The mothers obstetric and health history was examined, and indications revealed a generally low HB check at delivery and during postnatal care clinics but high HB checks during pregnancy at antenatal clinics. Usage of long-lasting insecticide bed nets was generally high (93.1%), with relatively low episodes of malaria infections after delivery (8.6%). Lastly, it was also found that iron supplements taken before delivery was seen to be high in pregnant women but low in lactating mothers during breastfeeding (16.0%). The majority (98.1: 412) of the lactating mothers had never dewormed after delivery of index child.

Also, the study identified 56.0% [95% **CI**: 51.25, 60.75%] of the lactating mothers living in the Nanton district of the northern region of Ghana to be anaemic; out of the total, 18.7% of the mothers were suffering from mild anaemia, 67.2% were moderately anaemic, and 14.1% were severely anaemic with 6.30g/dl as minimum haemoglobin level recorded and 14.8g/dl as maximum. These findings depict one of the highest among developing countries in the world as India reported 66.0% of lactating mothers had been reported having anaemia [24], Kenya reported 43.8% of lactating mothers with a HB level below 12.0 g/dL [25], and Myanmar reported an anaemia prevalence rate of 60.3% in breastfeeding mothers, with 20.3% diagnosed of severe anaemia [11]. More so, the findings of this study are unconnected with the study conducted in Ethiopia, where the overall prevalence of anaemia among lactating mothers from 2005–to 2011 was 22.1%. The prevalence of anaemia for the years 2005 and 2011 was 29.9% and 18.5%, respectively. In the period 2005–2011, the highest prevalence of anaemia among lactating mothers was 48.7% found in the Somali region, while the lowest prevalence was 9.0% reported in Addis Ababa [23].

Statistical difference between the mean of anaemic and non-anaemic lactating mothers showed statistically significant difference (**F-stat**=68.233, **t-stat**=-35.697, **p**<0.01), indicating a true deviation and magnitude of anaemia amongst mothers diagnosed of anaemia which predominantly high than findings from Lakew, Biadgilign, and Haile (2015) who found that the odds of working lactating mothers being anaemic was 29% less than their counterparts (**AOR**=0.71; 95% **CI**: 0.63, 0.80). Lactating mothers with a normal maternal BMI (18.5–24.99 kg/m^2^) were 22% less likely to be anaemic than those with low BMI (<18.5 kg/m^2^) (**AOR**=0.78; 95% **CI**: 0.68, 0.89) [23].

The odds of having anaemia increased by 94.6% amongst lactating mothers who have had episode(s) of malaria after birth [**AOR** = 0.054; (95% **CI**: 0.025, 0.119)] compared to lactating mothers who did not have any episode(s) of malaria. This finding correlates with Berhanu and Teferi (2018) whose findings from a comparative study revealed that malaria-infected women had 3.61 folds higher risk of anemia as compared to women with no history of malaria infection [**AOR**=3.6; (95% **CI**: 2.63, 4.95)], as well as finding from north Ethiopia [27], Nepal [28], and Ghana [29]. This is because Plasmodium species ingests the host’s red blood cells and finally decreases the number of red blood cells [26].

Similarly, iron supplement intake during lactation decreases the risk of anaemia by 15.336 folds [**AOR** = 15.336; (95% **CI**: 6.112, 38.483)] more than lactating mothers who did not take iron supplements. These findings concord with studies that revealed that food fortification, food diversification, iron supplementation, nutrition education, and quality maternal health service provision are the main effective interventions recommended preventing micronutrient deficiencies and associated mortality among pregnant and lactating women in low-and middle-income countries [30, 31].

Lastly, lactating mothers had a higher risk [**AOR** = 1.927, (95% **CI**: 1.031, 3.602)] of developing anaemia with increasing ‘child’s age greater than three (3) months. This findings is in bifurcation with many other studies which found that ‘mother’s age remains a risk factor for anaemia during pregnancy; the higher the age, the higher the risk of developing anaemia [2], [30], [32].

### Conclusion

Anaemia is highly prevalent among lactating mothers, particularly mothers who do not take an iron supplement during lactation, who have had episode(s) of malaria episodes after birth, and mothers with children greater than three (3) months. It is therefore recommended that the Ghana Health Service should extend the iron supplement distribution and Ipt program for pregnant women to cover the PNC and CWC units of the health facilities as well as roll out interventions and special initiatives towards mitigating the findings of the study.

## Supporting information

Ethical Clearance

## Data Availability

The data for this study are readily available and upon request to the authors will be made available

## Acknowledgment

The authors of this paper wish to acknowledge the Ghana Health Service, Northern Regional Health Directorate, and Nanton District Health Directorate for granting access, permission, and support during the collection of primary and secondary data for the study.

## Ethics Approval

The permission and ethical approval for the study were granted by Ghana Health Service and Kwame Nkrumah University of Science and Technology ethics committee, respectively.

## ‘Authors’ contributions

The authors of the research contributed in diverse areas with specific roles and responsible as denoted;

Adadow Yidana, contributed to the manuscript write-up and analysis

Enoch Weikem Weyori, contributed in analysis of data and presentation of results and findings

Khadijah Wemah, contributed in the write up and funding of the research

Nkrumah Bernard, contributed to the manuscript review and funding for the research

Braimah Baba Abubakari, contributed to the manuscript review and funding

Kuugbee Dogkotenge Eugene, contributed to the manuscript write-up and review as well as analysis

Benjamin Nuertey^7^, contributed to manuscript review and funding

Etowi Boye Yakubu, contributed to analysis and write-up of the manuscript

Valentine Koyiri Cheba, sample collection and laboratory testing, and data management

Adatsi Richard Kujo, sample collection, and laboratory testing, and data management

## Data Sharing Statement

Despite receiving ethics approval from CHRPE and GHS, the data for this study will be shared because the data was gathered from respondents who gave their consent before participating in the study.

