## Supplementary material for "Enormity of anaemia and its determinant factors among lactating mothers in Northern Ghana: A case of nanton district": Ethical Clearance

KWAME NKRUMAH UNIVERSITY OF SCIENCE AND TECHNOLOGY
**COLLEGE OF HEALTH SCIENCES**
SCHOOL OF MEDICAL SCIENCES / KOMFO ANOKYE TEACHING HOSPITAL
***COMMITTEE ON HUMAN RESEARCH, PUBLICATION AND ETHICS***

Our Ref: CHRPE/AP/102/21

10th March, 2021.

Ms. Amadu Wemah Khadijah
University for Development Studies
TAMALE.

Dear Madam,

**LETTER OF APPROVAL**

***Protocol Title:*** ***"Assessing the Knowledge, Prevalence and Factors Influencing the Anaemia Status of Lactating Mothers in the Nanton District."***

***Proposed Site:*** ***Health Facilities, Nanton District.***

***Sponsor:*** ***Principal Investigator.***

Your submission to the Committee on Human Research, Publications and Ethics on the above-named protocol refers.

The Committee reviewed the following documents:

- A Completed CHRPE Application Form.
- Participant Information Leaflet and Consent Form.
- Research Protocol.
- Questionnaire.

The Committee has considered the ethical merit of your submission and approved the protocol. The approval is for a fixed period of one year, beginning **10th March, 2021** to **9th March, 2022** renewable thereafter. The Committee may however, suspend or withdraw ethical approval at any time if your study is found to contravene the approved protocol.

Data gathered for the study should be used for the approved purposes only. Permission should be sought from the Committee if any amendment to the protocol or use, other than submitted, is made of your research data.

The Committee should be notified of the actual start date of the project and would expect a report on your study, annually or at the close of the project, whichever one comes first. It should also be informed of any publication arising from the study.

Thank you, Madam, for your application.

Yours faithfully,

Rev. Prof. John Appiah-Poku.
**Honorary Secretary**
**FOR: CHAIRMAN**

Room 7 Block J, School of Medical Sciences, KNUST, University Post Office, Kumasi, Ghana
